# Towards Assessing Subcortical “Deep Brain” Biomarkers of PTSD with Functional Near-Infrared Spectroscopy

**DOI:** 10.1101/2022.06.03.22275966

**Authors:** Stephanie Balters, Marc R. Schlichting, Lara Foland-Ross, Sabrina Brigadoi, Jonas G. Miller, Mykel J. Kochenderfer, Amy S. Garrett, Allan L. Reiss

## Abstract

Assessment of brain function with functional near-infrared spectroscopy (fNIRS) is limited to the outer regions of the cortex. Previously, we demonstrated the feasibility of inferring activity in subcortical “deep brain” regions using cortical fMRI and fNIRS activity in healthy adults. Access to subcortical regions subserving emotion and arousal using affordable and portable fNIRS is likely to be transformative for clinical diagnostic and treatment planning. Here, we validate the feasibility of inferring activity in subcortical regions that are central to the pathophysiology of PTSD (i.e., amygdala and hippocampus) using cortical fMRI and simulated fNIRS activity in a sample of adolescents diagnosed with PTSD (*N*=20, mean age=15.3±1.9 years) and age-matched healthy controls (*N*=20, mean age=14.5±2.0 years) as they performed a facial expression task. We tested different prediction models, including linear regression, a multi-layer perceptron neural network, and a k-nearest neighbors model. Inference of subcortical fMRI activity with cortical fMRI showed high prediction performance for the amygdala (*r>*0.91) and hippocampus (*r>*0.95) in both groups. Using fNIRS simulated data, relatively high prediction performance for deep brain regions was maintained in healthy controls (*r>*0.79), as well as in youths with PTSD (*r>*0.75). The linear regression and neural network models provided the best predictions.

## 1 Introduction

At least two thirds of youths within the United States are exposed to a traumatic event by late adolescence (Giaconia et al. 1995; Copeland et al. 2007). As a result, many of these youths are at risk of developing Post-Traumatic Stress Disorder (PTSD) (Macdonald et al. 2010; Alisic et al. 2014). Traumatic and Adverse Childhood Experiences (TRACEs) have been associated with a number of psychological and behavioral difficulties including anxiety, hypervigilance, aggression, flashbacks, nightmares, avoidance of situations that induce trauma memories, sleep disturbance, and impaired cognitive and executive functioning (Yoon et al. 2016; Aebi et al. 2017; Malarbi et al. 2017; Weems, Russell, et al. 2021; Weems, McCurdy, et al. 2021). In addition, PTSD in early life is associated with social and academic impairments that have long-term implications for negative outcomes in adulthood (Cohen et al. 2018).

Findings from clinical neuroimaging show the potential of utilizing neural signatures as brain biomarkers of PTSD. Using functional Magnetic Resonance Imaging (fMRI) data, researchers have consistently achieved differentiation of adults with PTSD from healthy controls (Rabinak et al. 2011; Chen et al. 2018), as well as prediction of PTSD symptom severity (Gong et al. 2014; Wang et al. 2016; Stevens et al. 2017; Misaki et al. 2018; Kim et al. 2019; Fitzgerald et al. 2020), symptom improvement (Fonzo et al. 2017; Joshi et al. 2020; Bryant et al. 2021), and even PTSD pathogenesis (Belleau et al. 2020; Sheynin et al. 2021). Less research has been conducted on pediatric PTSD. However, results of the few available studies suggest the neural hallmarks of this condition are in line with those of adult PTSD and demonstrate the feasibility of differentiating youths with PTSD from healthy controls (Garrett et al. 2012; Keding and Herringa 2016; Cohen et al. 2018), and predicting PTSD severity (Carrion et al. 2010) and PTSD symptom improvement in youth cohorts (Cisler et al. 2016; Garrett et al. 2019). In most of the extant PTSD neuroimaging literature, subcortical “deep brain” regions, such as the amygdala and hippocampus, are found to serve as key regions for the establishment of predictive associations with PTSD symptomatology in adults (Fonzo et al. 2017; Stevens et al. 2017; Misaki et al. 2018; Kim et al. 2019; Belleau et al. 2020; Fitzgerald et al. 2020; Kaldewaij et al. 2021) and youths (Garrett et al. 2012; Cisler et al. 2016; Keding and Herringa 2016; Garrett et al. 2019). Both the amygdala and hippocampus are integral parts of the limbic system and involved in emotion formation, processing, and regulation (Banks et al. 2007; Shackman et al. 2011; Stevens et al. 2011). Their aberrant functioning is thought to contribute to the hypervigilance and the heightened threat and fear response in PTSD (Sripada et al. 2012; Koch et al. 2016; Herringa 2017; Harnett et al. 2020; Zhang et al. 2021; Alexandra Kredlow et al. 2022) – all key symptoms in PTSD (American Psychiatric Association et al. 2013). Despite the enormous potential of utilizing fMRI for PTSD brain biomarker detection, its high cost and cumbersome environmental requirements are major limitations that impede frequent usage in the applied clinical context. Functional MRI is currently the only neuroimaging technique that allows for the functional assessment of these subcortical brain biomarkers.

In research aimed at overcoming the financial and methodological challenges of assessing subcortical brain function, our laboratory has demonstrated the feasibility of inferring fMRI subcortical activity using cortical signals from functional near-infrared spectroscopy (fNIRS) (Liu et al. 2015). Functional NIRS is a portable and relatively affordable neuroimaging technology with relatively high spatial resolution in cortical areas (Cui et al. 2011; Scholkmann et al. 2014), allowing researchers to assess dynamic changes in oxygenation across the surface of the brain, typically with 2-3cm measurement depth in adults (Cui et al. 2011).^1^ Many deep brain regions are anatomically and functionally connected with cortical surface regions (Bullmore and Sporns 2009; Power et al. 2011). The deep brain inference method leverages this connectivity to infer deep brain activity through cortical surface activity patterns that are accessible to fNIRS.

In a previous study from our group, 17 healthy adults engaged in a facial expression task while being scanned with concurrent fMRI-fNIRS. Support vector regression was applied to infer deep brain activity (e.g., amygdala and hippocampus) on the basis of cortical fMRI (i.e., ground truth) and fNIRS cortical activity in the prefrontal cortex (PFC). Average prediction performance across deep brain regions was high (*r*=0.67) for cortical fMRI input and the top 15% of predictions using fNIRS signals achieved an accuracy of *r*=0.70 (Liu et al. 2015). Extending the deep brain inference method to the study of PTSD has important clinical implications. Access to subcortical biomarkers (i.e., amygdala and hippocampus activity) with affordable and portable fNIRS could inform clinical diagnostic and treatment planning. Additionally, researchers could study deep brain activity and associated PTSD behavior (e.g., heightened fear, anxiety, and aggression responses) in naturalistic settings, which would advance understanding of brain mechanisms underlying PTSD in youths and adults. However, because PTSD is associated with aberrant brain function in both deep brain and NIRS-inaccessible cortical regions (Cui et al. 2011; Suarez-Jimenez et al. 2020; Bao et al. 2021), it is unclear whether the deep brain inference method is suitable for individuals with PTSD. Additionally, changes in functional connectivity related to brain development (Blakemore et al. 2010; Goddings et al. 2014; Vijayakumar et al. 2018) could pose further challenges for using this deep brain inference method in pediatric samples.

In this study, we validate the deep brain inference method in youths with PTSD and age-matched healthy controls. Specifically, we demonstrate the feasibility of inferring activity within two key subcortical regions in PTSD – the amygdala and hippocampus – based on cortical fMRI and simulated fNIRS data. Although our primary aim is to test the feasibility of the deep brain inference method for simulated fNIRS data, we also conduct inference predictions with cortical fMRI data to generate a performance “gold standard” benchmark. For this proof-of-concept study, we utilized an existing fMRI dataset [29]. Youths with PTSD (*N*=20) and healthy controls (*N*=20) were scanned using fMRI as they engaged in a facial expression task that reliably activates emotion regulation circuits (Carrion et al. 2010; Cisler et al. 2016; Garrett et al. 2019; Balters et al. 2021). We extracted cortical and subcortical activity from regions of interest defined by anatomical atlases (Frazier et al. 2005; Desikan et al. 2006; Makris et al. 2006; Goldstein et al. 2007). We analyzed these data in conjunction with Monte Carlo simulations of photon transport in spatially overlapping and anatomically informed multi-layer models (Brigadoi and Cooper 2015) to build a structural fNIRS cortical sensitivity map. We used this map to compute regional estimates of fNIRS activity based on the available fMRI data (i.e., “simulated fNIRS activity”). We hypothesized that cortical fMRI and simulated fNIRS data can be used to accurately infer bilateral amygdala and bilateral hippocampus activity in youths with and without PTSD. We tested different prediction approaches, including linear regression (with and without rolling window), a multi-layer perceptron (MLP) neural network, and a k-nearest neighbors (KNN) model, to allow us to (1) identify the best performing model; and, (2) learn more about the underlying structure of the problem (i.e., level of linearity, effect of measurement noise, and time-dependency).

## 2 Materials and Methods

### 2.1 Participants

We conducted the current analysis using data that were previously published by Garrett and colleagues (Garrett et al. 2019). The original study examined longitudinal changes in brain function associated with symptom improvement in youths with PTSD and was approved by the Stanford University Institutional Review Board. Written consent was obtained from all caregivers and youths. A total of *N*=20 youths with PTSD (18 females, 2 males, mean age: 15.3 ± 1.9 years, age range: 10.4 - 17.7 years) and *N*=20 age-, sex- and IQ-matched healthy controls (18 females, 2 males, mean age: 14.5 ± 2.0 years, age range: 11.2 - 17.5) participated in the study. All participants were post-onset of puberty based on self-reported Tanner Stages of 2 or above (Morris and Udry 1980). Individuals in the PTSD group met DSM-IV criteria for PTSD at baseline (i.e., pre-treatment) as assessed by *Schedule for Affective Disorders and Schizophrenia for School-Age Children-Present and Lifetime Version* (KSADS-PL) interviews (Kaufman et al. 1997). Based on the UCLA PTSD Reaction Index for DSM-IV (average of parent and child scores; (Steinberg et al. 2013)), the average self-reported PTSD symptoms score was 39.1 ± 10.6, indicating moderate symptom severity. Index trauma were interpersonal in nature for all individuals with PTSD. Fifty percent of the PTSD group met criteria for co-morbid major depression based on KSADS-PL interviews (Kaufman et al. 1997). None of the participants in either group were taking psychotropic medication or had any learning disability, neurological disorders, traumatic brain injury, major medical illness, an IQ *<* 70, or were currently hospitalized.

### 2.2 MRI Data Acquisition

Participants engaged in a widely used facial expression task, including photographs of happy, angry, and neutral faces. Fearful and angry faces have been extensively used to elicit emotion regulation responses (Sabatinelli et al. 2011). In addition, a growing body of research on pediatric PTSD suggests that neutral faces elicit activation in regions important for threat processing (Garrett et al. 2012; Keding and Herringa 2016; Garrett et al. 2019; Balters et al. 2021). Scrambled images were used as the contrasting condition. The task lasted a total of 8.5 minutes. In blocks of eight stimuli per condition, each face or scrambled image was presented for 3 seconds. Each block was repeated four times in pseudo-randomized order. Consistent with an implicit emotion task, participants were instructed to press button “1” for photographs of female, and button “2” for male faces. In the scrambled faces condition, participants alternated between button presses of “1” and “2” .

MRI data were collected on a 3 Tesla MR750 General Electric magnet (gehealthcare.com) using an 8-channel head coil. A spiral in/out pulse sequence was used to optimize signal to noise ratio while minimizing susceptibility artifacts (Lai and Glover 1998; Glover and Law 2001). Thirty axial slices (3mm thick, 1 mm skip), covering the entire brain were collected with the following parameters: field of view=24 cm, matrix=64x64; inplane resolution=3.125 mm; TR=2 seconds, TE=30 milliseconds; flip angle=70 degrees; 1 interleave. An individually calculated high-order shim for spiral acquisitions was used to reduce field heterogeneity (Lee et al. 2009).

### 2.3 Functional MRI Data Processing

We discarded the first three volumes of each scan to allow for magnet stabilization. Preprocessing was conducted using FSL software (https://fsl.fmrib.ox.ac.uk/fsl/fslwiki). Using the Brain Extraction Tool (BET), we processed structural and functional images to remove non-brain tissue (Smith 2002). We conducted additional motion correction to the mean image using MCFLIRT (Jenkinson et al. 2002), spatial smoothing with a Gaussian kernel of 6mm FWHM, and high-pass temporal filtering with a cutoff of 100s. We then aligned each subject’s functional image to their structural image using FMRIB’s Linear Image Registration Tool (Jenkinson and Smith 2001). With a linear registration, we aligned each subject’s structural intermediate image to a standard MNI template with an isotropic voxel size of 2mm (Fonov et al. 2011). We combined the linear transformations to register each subject’s functional data to MNI template space.

We conducted voxel-wise timeseries analyses separately for each participant. Specifically, we extracted the time course of activation from 48 discrete cortical regions, separated by hemisphere using the Harvard/Oxford atlas (Frazier et al. 2005; Desikan et al. 2006; Makris et al. 2006; Goldstein et al. 2007). This resulted in 96 regions for the cortical fMRI data. Using this same atlas, we extracted amygdala and hippocampus separately from both hemispheres, resulting in a total of four discrete deep brain regions. Given the probabilistic nature of the Harvard/Oxford atlases, we further eliminated any overlapping voxels between subcortical and cortical regions. Specifically, we built the union of all four deep brain regions, binarized and inverted this new mask, and multiplied it with each of the 96 fMRI cortical maps individually.

### 2.4 Simulation of fNIRS Data

As described earlier, we utilized a structural fNIRS cortical sensitivity map to compute regional estimates of fNIRS activity based on the available functional MRI activity data (i.e., “simulated fNIRS activity”). To build the structural fNIRS cortical sensitivity map, we applied Monte Carlo simulations of photon transport in anatomically informed multi-layer models following similar procedures described in Brigadoi and Cooper (2015). We built the model from the nonlinear symmetric MNI-ICBM152 atlas template (Fonov et al. 2011). We obtained the multi-layer tissue mask, segmented into five tissue types (scalp, skull, cerebrospinal fluid, grey matter, and white matter) using the MRI tissue probability maps for brain tissue segmentation and FSL betsurf (https://fsl.fmrib.ox.ac.uk/fsl/fslwiki/BET/) for scalp and skull segmentation. We further eroded the outer scalp layer with a 6-pixel cube due to overestimation of the scalp layer by FSL (Perdue and Diamond 2014) mainly caused by the blurring of the T1-weighted image at the border between scalp and air. We created the tetrahedral volumetric mesh using the cgalmesher option in the iso2mesh toolbox (Fang and Boas 2009). We set the maximum element volume to 8mm^3^ and the maximum radius of the Delaunay sphere to 1.5mm. The head model is freely available online (https://github.com/DOT-HUB/ArrayDesigner). We calculated the 10-5 positions of the international EEG system (Oostenveld and Praamstra 2001) on the outer scalp surface, based on manually identified cranial landmarks (nasion, inion, pre-auricolar points), following the procedure described in Brigadoi and Cooper (2015). As shown in **Figure 4** in the Appendix, we used the 10-5 positions as possible locations for fNIRS sources/detectors that would provide extensive coverage of the cortex. We estimated the sensitivity of simulated optodes located in the 10-5 positions using an approach similar to the one proposed in Brigadoi et al. (2018). For each 10-5 position, we performed photon migration simulations with the TOAST++ software (Schweiger and Arridge 2014). We assigned optical properties to each tissue type by fitting all published adult values across the near-infrared spectrum and selecting the fitted values at the 800nm wavelength (Bevilacqua et al. 1999; Strangman et al. 2003; Custo et al. 2006). We therefore computed fluence distributions for each 10-5 positions in the volumetric head model mesh. Next, we computed photon measurement density functions (PMDF) for each viable channel (any possible 10-5 pair with a maximum distance of 45mm) using the adjoint method. This method multiplies the fluence distribution of the source with the fluence distribution of the detector, correcting for the nodal volume, and normalizing by the value of the fluence distribution at the source/detector position (the largest one). We then computed the array sensitivity by summing the PMDFs of all viable channels. Channels were weighted by signal-to-noise ratios estimated from source-detector distance (i.e., signal-to-noise decreases as source-detector distance increases (Brigadoi et al. 2018). We computed this factor considering a maximum allowed source-detector distance of 45mm, with a maximum source-detector distance allowing to measure reliable data with good signal-to-noise ratio of 35mm. We further restricted the sensitivity matrix to the grey matter volumetric mesh and thresholded so that all values lower than a coverage threshold were set to zero. To define the coverage threshold, or minimum sensitivity at which the array is considered to effectively measure from that area, we used methods described in Brigadoi et al. (2018). Specifically, we considered a node to be covered when a change in absorption coefficient by 0.0001mm^2^ in a block of tissue of 0.5cm^3^ yielded a change in intensity in the measured fNIRS signal larger than 1%. We converted the final thresholded sensitivity matrix to the original voxel space of the MNI152 atlas by assigning to each voxel the mean sensitivity value of all nodes within a 2mm distance from the center of the voxel in all 3D directions (Figure 1). Lastly, we utilized this structural fNIRS cortical sensitivity map to compute estimates of fNIRS activity based on the available Harvard-Oxford cortical MRI estimates to produce voxel-based “simulated” fNIRS timeseries. The feature space consists of 58 cortical regions for simulated fNIRS data.

**Figure 1:**
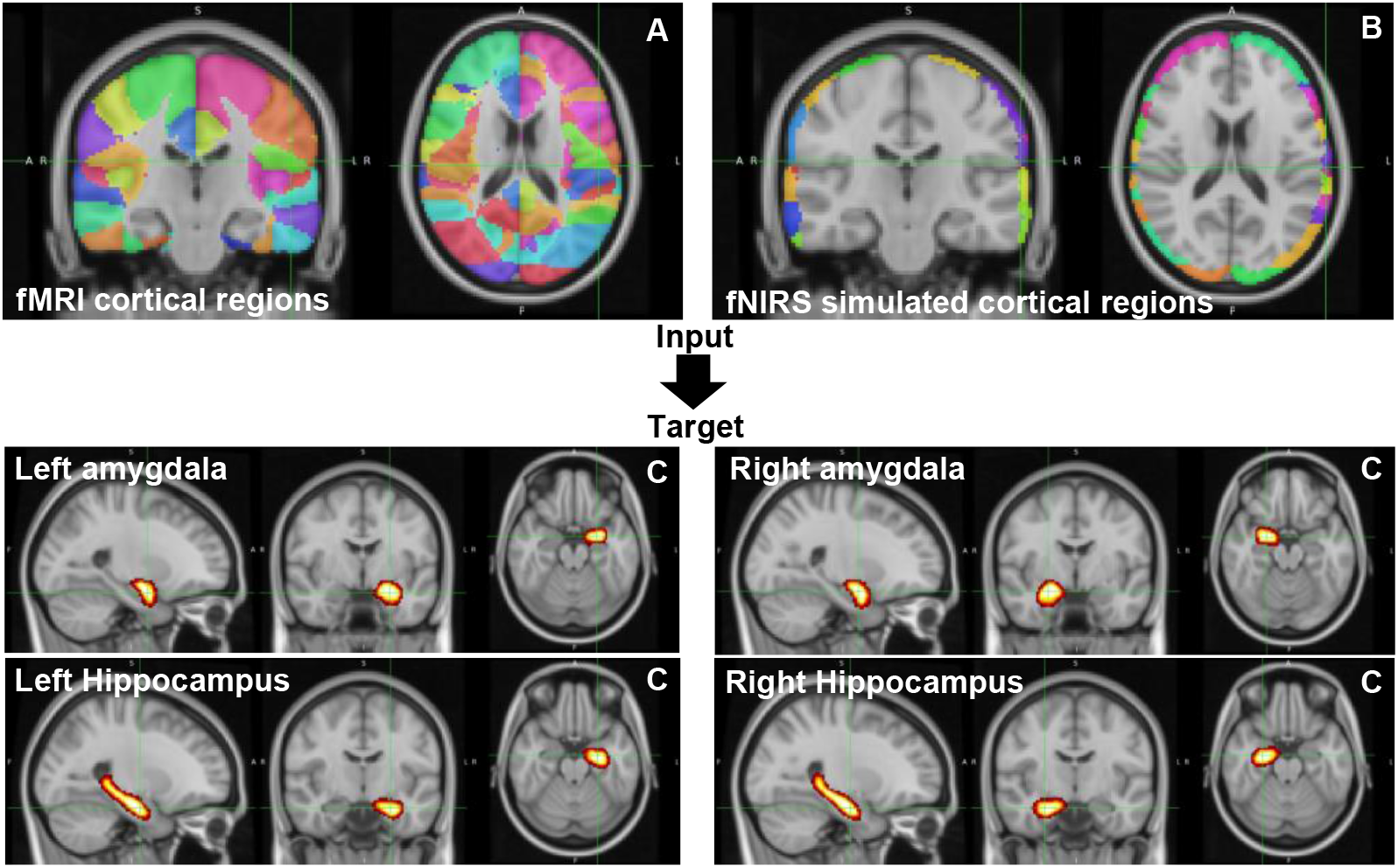
Overview of the Deep Brain Inference Method. We tested the feasibility of inferring deep brain activity in subcortical “deep brain” regions based on cortical fMRI (ground-truth) and simulated fNIRS data. Specifically, we used fMRI cortical data from 96 cortical regions as defined by the Harvard-Oxford Atlas (**A**). We further built a fNIRS sensitivity map via Monte Carlo simulation of photon transport. We utilized this sensitivity map to compute regional estimates of fNIRS activity based on the available functional MRI activity data (i.e., “simulated fNIRS activity”) (**B**). As deep brain target regions, we chose the bilateral amygdala and bilateral hippocampus because both are central to the pathophysiology of PTSD (**C**).

### 2.5 Deep Brain Inference Method

For the deep brain inference method, we used three different data streams. As target data, we used the deep brain activity of the bilateral amygdala and bilateral hippocampus (i.e., four deep brain regions). As input data, we used the activity of 96 cortical fMRI regions as well as the activity of 58 cortical simulated fNIRS regions. To ensure the comparability between different runs and different participants, we normalized the time signals of all three data streams. We conducted prediction analyses for the two groups separately (i.e., youths with and without PTSD). For each group, we split the dataset into a training dataset consisting of 70% of the subjects and a test dataset containing the remaining 30% of subjects. Splitting the dataset subject-wise and not data-point wise increases the likelihood that our models generalize across different subjects within the same group.

We compared four different machine learning models to explore which best predicts activity of deep brain regions (i.e., method that produces predicted activity values that have the highest Pearson correlation with observed activity values). The four models that we tested were linear regression with L2-regularization (Russell and Norvig 2021), linear regression with rolling windows (Hota et al. 2017), a multi-layer perceptron neural network with dropout (Srivastava et al. 2014), and a k-nearest neighbor model (Fix and Hodges 1989; Altman 1992).

#### 2.5.1 Linear Regression

In linear regression, a hyperplane which is linear in the features is fitted using a least-squares cost function. Using the negative correlation coefficient as a cost function works as well, but we found that using the ordinary least-squares cost function yields better results in terms of the correlation coefficient. To avoid overfitting due to the limited size of the dataset, we use L2-regularization, which appends a term to the cost function that is based on the sum of squares of the weights. The prediction function in terms of the design matrix *X* is:

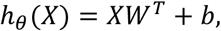

where *W* ∈ ℝ^*m*×*d*^ are the weights and *b* ∈ ℝ^*m*^ is the bias term. The cost function is:

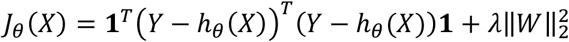

where **1** denotes a column vector of ones and *λ* is the regularization parameter.

Using the cost function, gradient descent is used to change the parameters *θ* in order to minimize the cost function *J*. We use the Adam optimizer for this task (Kingma and Ba 2014).

#### 2.5.2 Linear Regression with Rolling Time Windows

So far, it was assumed that all the data points are independent of each other; however, since the goal is to analyze a time series, this cannot be assumed by default. Thus, the features for each of the cortical regions are augmented by time-wise previous values for the same region. For example, instead of only using the measurement of the bilateral dorsolateral prefrontal cortex at time *t*, the measurements up to *t* − *p*, where *p* + 1 is the window length, are also included as a feature. The same strategy as for linear regression is used, but the design matrix then becomes *X* ∈ ℝ^*n*×*pd*^, and the weight becomes *W* ∈ ℝ^*m*×*pd*^.

#### 2.5.3 Multi-Layer Perceptron Neural Network

We used a *multi-layer perceptron* (MLP), which is a simple form of a neural network and is an extension of the linear model described in section 2.5.1. There are two significant differences concerning the model architecture that distinguish the MLP from a linear model. As the name suggests, there are multiple layers that are sequentially concatenated, where the output of one layer is the input to the next layer. Secondly, to make the model non-linear, a non-linearity *g* is applied to each output of the linear layer with exception to the last layer to provide more flexibility to the output. Because our data is normalized, we decided to use a hyperbolic tangent as non-linearity. The non-linearity is important to make the model more expressive, i.e., enabling the model to learn complex, non-linear relationships between the features and the targets (Russell and Norvig 2021). Each layer’s output can be described as:

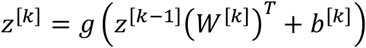

where *W*^[*k*]^ is the weight matrix and *b*^[*k*]^ as bias vector of the same size as *z*^[*k*]^. All intermediate values of *z*^[*k*]^ that are not the output are referred to as hidden layers (with *k* as enumerator) and the dimension of each state is colloquially referred to as the number of neurons. For our tests, we use only one hidden layer of the same size as the input which still allows for more expressiveness of the model than a linear model. For training, gradient-based methods are used because no closed-form solution exist. A typical choice is the Adam optimizer because it outperforms many other optimizers and delivers robust performance (Kingma and Ba 2014). To prevent overfitting, the weights are L2-regularized. We also apply a dropout layer to the hidden layer to make the model more robust to noise in the training dataset (Srivastava et al. 2014).

#### 2.5.4 K-Nearest Neighbors Model

The *K-Nearest Neighbors* (KNN) model is a parameter-free model as it only keeps the training data to make predictions and does not fit parameters. Inherently, it is a non-linear model, as for each input the output is calculated as the mean of the outputs of the k-nearest neighbors in the training set (Fix 1985; Altman 1992). The prediction can be formalized by assuming a set *S* which consist of pairs (*x*^(*i*)^, *y*^(*i*)^). Let 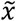 be the feature in question for the prediction. The prediction can be obtained by ordering *S* in such a way that 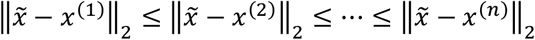. Assuming such an ordered set, the prediction can be obtained by:

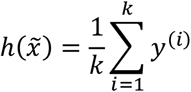

where *y*^(*i*)^ is corresponding target to *x*^(*i*)^. The value of *K* is a hyper parameter that we can optimize during training.

### 2.6 Assessment of Prediction Performance

To train the four parameter-based models, we minimize mean squared error. We also use the Pearson’s correlation coefficient (*r*) to measure the degree of association between the target and predicted values (Student 1908). The higher the *r*-value, the better the performance of the prediction model. Because of the stochastic nature of the parameter initialization and training for the linear regression and multi-layer perceptron, we train each model with 10 different random seeds, which is common practice in machine learning studies (Henderson et al. 2018). We report the mean and standard deviation from those experiments to show that our methods are stable and converge to similar results independently of the random seeds. Because we contrast prediction results across different models, groups, and data inputs, we only report *r*-values that pass false discovery rate (FDR) correction (*p<*0.05).

To contrast the prediction performances of different models, we conducted additional statistical tests. We transformed all *r*-values (i.e., 10 *r*-values for each test) to Fisher z-statistics. First, we performed four different analyses of variance (ANOVAs) to assess the best prediction model for each prediction case across all deep brain regions. To account for multiple testing, we FDR-corrected the resulting *p*-values for four comparisons. Next, we conducted a two-way ANOVA to determine the effect of input data (i.e., cortical fMRI versus simulated fNIRS) and/or group (i.e., healthy controls versus youths with PTSD) on the observed performance outcomes. We then followed up with a series of paired samples t-test to gain more detailed information about the underlying differences (i.e., separate group differences analyses for cortical fMRI and simulated fNIRS data; and separate data input differences analyses for the healthy controls group and PTSD group). We FDR-corrected across all t-tests.

## 3 Results

All Pearson’s correlations tests were statistically significant and passed FDR-correction for multiple testing (all FDR-corrected *p*<0.001). In **Table 1**, we present the prediction results for each model prediction. Because we conducted each prediction ten times (i.e., with ten random seeds), we noted the mean values and standard deviation across these ten predictions.

**Table 1:** Performance results for all prediction models. All Pearson’s correlations tests were statistically significant and passed FDR-correction for multiple testing (all FDR-corrected *p*<0.001). We tested the four different prediction models for each case, including using cortical fMRI data as input data for prediction in the healthy control group (**case 1**) or the PTSD group (**case 2**), and using simulated fNIRS data as input data for prediction in the healthy control group (**case 3**) or the PTSD group (**case 4**). The highest correlation coefficient for each case is presented in bold font. We tested models involving randomness in the training process (i.e., linear regression with and without rolling window and MLP) with 10 random seeds. We noted mean values and standard deviations across these 10 tests. For deterministic models (i.e., KNN), we do not report a standard deviation because the deterministic model creates the same output ten times. Abbreviations: linear regression with rolling window = LR w/roll. window, multi-layer perceptron = MLP, and k-nearest neighbors = KKN.

|  | Input Data | Cohort | Prediction Model | Correlation Coefficient |  |  |  |
| --- | --- | --- | --- | --- | --- | --- | --- |
|  |  |  |  | Left Amygdala | Right Amygdala | Left Hippocampus | Right Hippocampus |
| Case 1 | Cortical fMRI | Controls | **Linear regression** | **0.937±6\*10<sup>-6</sup>** | **0.912±9\*10<sup>-6</sup>** | **0.972±1\*10<sup>-7</sup>** | **0.956±9\*10<sup>-8</sup>** |
|  | Cortical fMRI | Controls | LR w/ roll. window | 0.867 ± 0.021 | 0.814 ± 2*10<sup>-4</sup> | 0.922 ± 3*10<sup>-4</sup> | 0.871 ± 5*10<sup>-5</sup> |
|  | Cortical fMRI | Controls | MLP | 0.932 ± 6*10<sup>-4</sup> | 0.909 ± 5*10<sup>-4</sup> | 0.969 ± 7*10<sup>-4</sup> | 0.953 ± 5*10<sup>-4</sup> |
|  | Cortical fMRI | Controls | KNN | 0.893 | 0.869 | 0.925 | 0.908 |
| Case 2 | Cortical fMRI | PTSD | **Linear regression** | **0.911±3\*10<sup>-7</sup>** | **0.913±2\*10<sup>-4</sup>** | **0.954±8\*10<sup>-6</sup>** | **0.949±4\*10<sup>-5</sup>** |
|  | Cortical fMRI | PTSD | LR w/ roll. window | 0.824 ± 0.009 | 0.799 ± 6*10<sup>-4</sup> | 0.861 ± 6*10<sup>-4</sup> | 0.859 ± 1*10<sup>-4</sup> |
|  | Cortical fMRI | PTSD | MLP | 0.910 ± 0.001 | 0.912 ± 0.001 | 0.952 ± 5*10<sup>-4</sup> | 0.948 ± 7*10<sup>-4</sup> |
|  | Cortical fMRI | PTSD | KNN | 0.839 | 0.854 | 0.888 | 0.898 |
| Case 3 | Simulated fNIRS | Controls | **Linear regression** | **0.812±9\*10<sup>-8</sup>** | **0.791±1\*10<sup>-8</sup>** | **0.854±6\*10<sup>-8</sup>** | **0.839±1\*10<sup>-6</sup>** |
|  | Simulated fNIRS | Controls | LR w/ roll. window | 0.719 ± 0.001 | 0.646 ± 0.002 | 0.778 ± 0.004 | 0.717 ± 3*10<sup>-5</sup> |
|  | Simulated fNIRS | Controls | MLP | 0.797 ± 0.002 | 0.789 ± 0.002 | 0.827 ± 0.002 | 0.822 ± 0.002 |
|  | Simulated fNIRS | Controls | KNN | 0.791 | 0.772 | 0.814 | 0.798 |
| Case 4 | Simulated fNIRS | PTSD | Linear regression | 0.724 ± 8*10<sup>-8</sup> | 0.737 ± 3*10<sup>-4</sup> | 0.749 ± 7*10<sup>-8</sup> | 0.787 ± 1*10<sup>-4</sup> |
|  | Simulated fNIRS | PTSD | LR w/ roll. window | 0.613 ± 1*10<sup>-4</sup> | 0.619 ± 0.002 | 0.695 ± 9*10<sup>-7</sup> | 0.699 ± 4*10<sup>-4</sup> |
|  | Simulated fNIRS | PTSD | **MLP** | **0.745±0.002** | **0.768±0.002** | **0.773±0.002** | **0.808±0.001** |
|  | Simulated fNIRS | PTSD | KNN | 0.713 | 0.749 | 0.768 | 0.797 |

### 3.1 Identification of Highest Predicting Model for Each Prediction Case 1-4

ANOVA results demonstrated statistically significant differences in prediction performance between the four prediction models in all of the four prediction cases (**case 1**: F(3,117) = 594.165, FDR-corrected *p*<0.001, partial η^2^ = 0.938; **case 2**: F(3,117) =1115.85, FDR-corrected *p*<0.001, partial η^2^ = 0.966; **case 3**: F(3,117) =521.01, FDR-corrected *p*<0.001, partial η^2^ = 0.930; **case 4**: F(3,117) =878.550, FDR-corrected *p*<0.001, partial η^2^ = 0.957; see **Table 1**). We conducted post-hoc analyses with FDR-correction. For cases 1-3, the linear regression without rolling window consistently had the highest prediction performance values (all FDR-corrected *p*<0.001, with mean differences to the other three models ranging between 0.008 and 0.482). In contrast, linear regression with rolling window had the lowest prediction performance for all three cases (all FDR-corrected *p*<0.001, with mean differences to the other three models ranging between -0.482 and -0.125). For case 4, the MLP neural network had the highest prediction performance (all FDR-corrected *p*<0.001, with the mean difference to the other three models ranging between 0.039 and 0.242), whereas linear regression with rolling window had the lowest prediction performance (all FDR-corrected p<0.001, with the mean difference to the other three models ranging between -0.183 and -0.204). Given that linear regression had the best prediction performance, we present the corresponding prediction weights for this model across all cortical fMRI regions and all simulated fNIRS regions in **Figure 2** and **Figure 3**, respectively. We use these cortical weight maps as additional descriptive information when interpreting the results.

**Figure 2:**
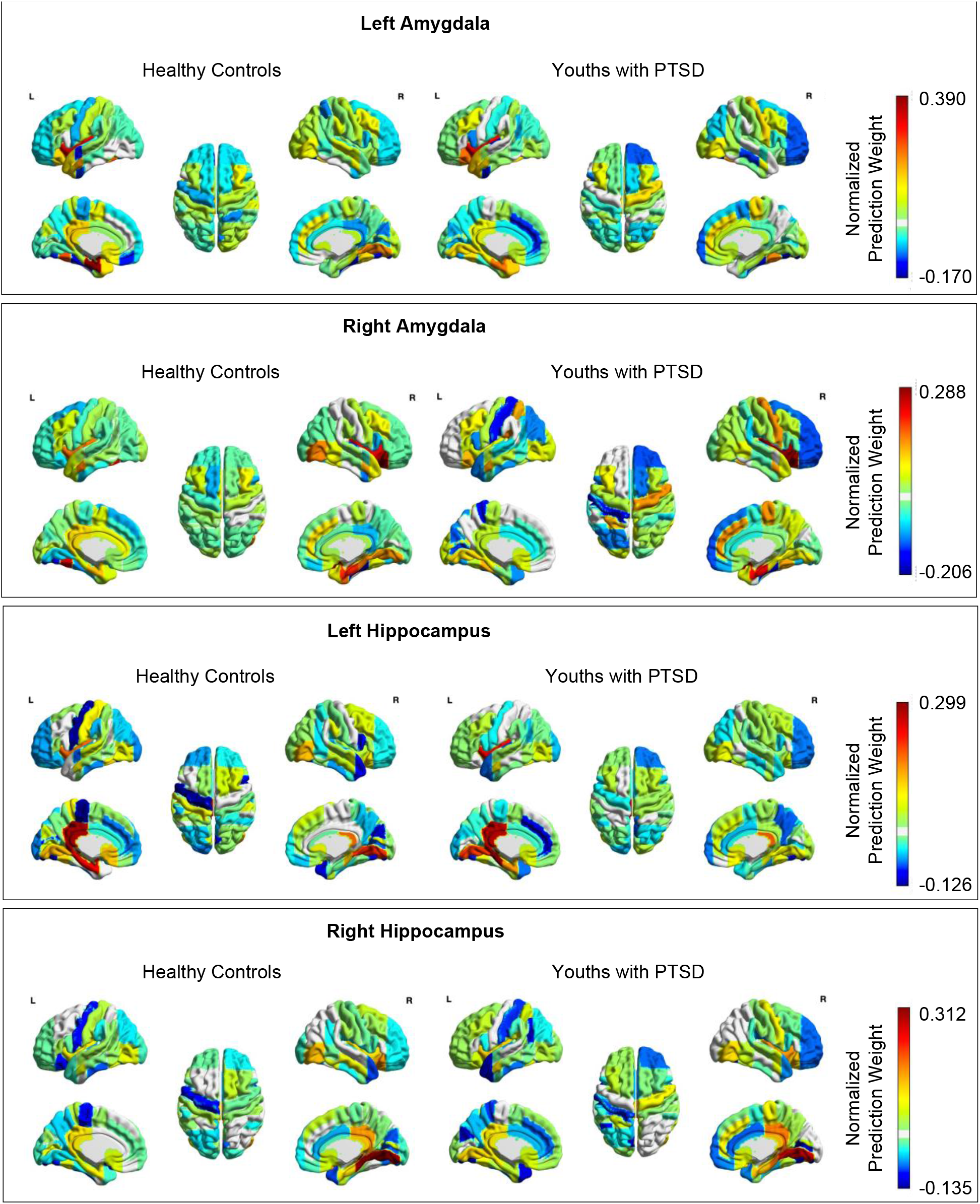
Cortical weight maps for the linear regression results of the cortical fMRI prediction. Positive values are represented by warmer colors and reflect a positive association between the activity of the cortical region defined by the Harvard/Oxford cortical atlas and the activity of the deep brain region. Negative values are represented by cooler colors and reflect a negative association between the activity of the cortical region defined by the Harvard/Oxford cortical atlas and the activity of the deep brain region.

**Figure 3:**
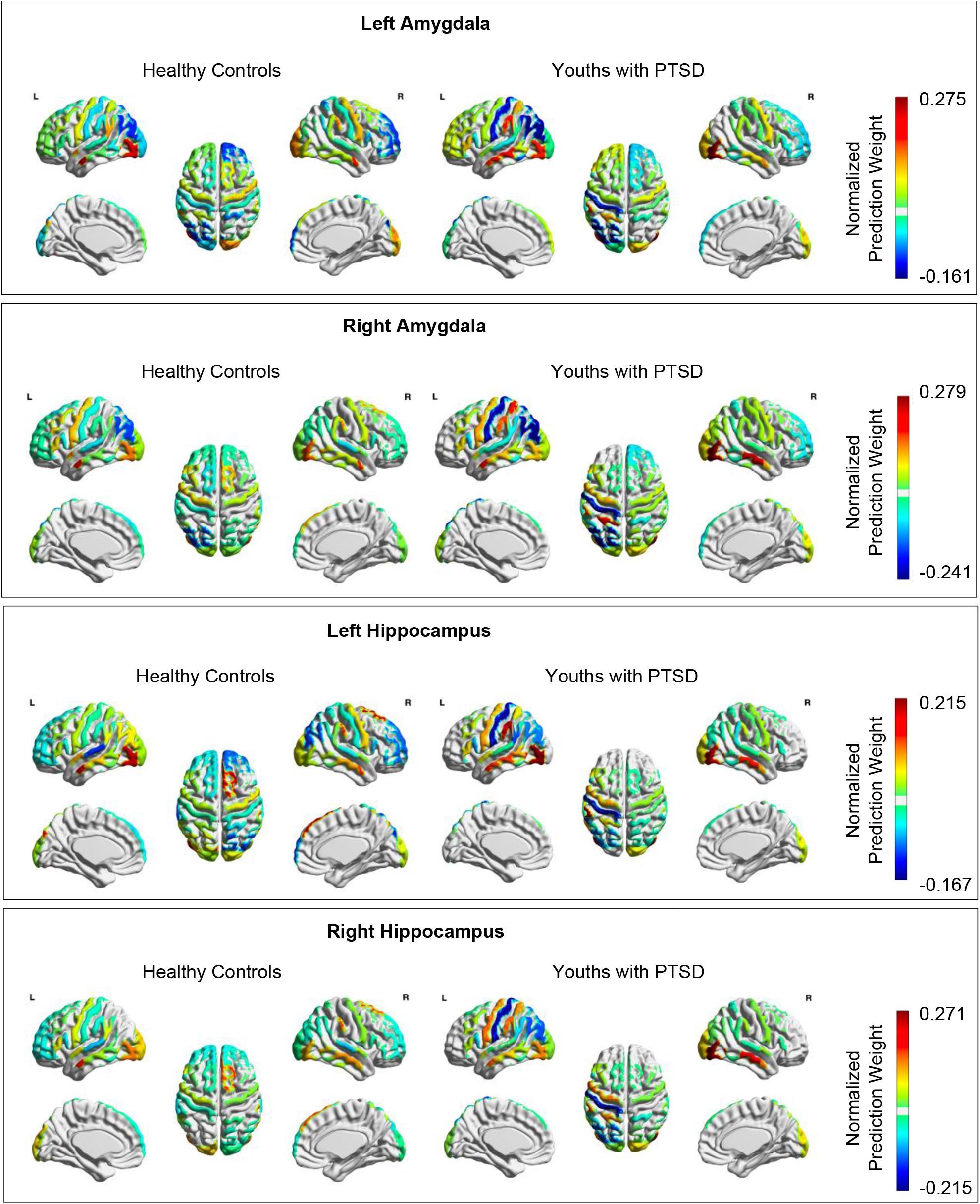
Cortical weight maps for the linear regression results of the simulated fNIRS prediction. Positive values are represented by warmer colors and reflect a positive association between the activity of the cortical region from the fNIRS simulation and the activity of the deep brain region. Negative values are represented by cooler colors and reflect a negative association between the activity of the cortical region from the fNIRS simulation and the activity of the deep brain region.

**Figure 4.**
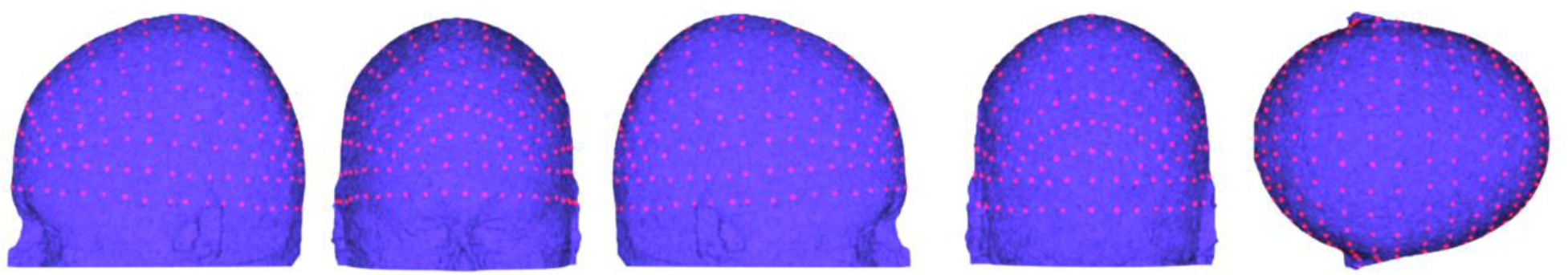
The 10-5 positions of the international EEG system [68].

### 3.2 Performance Differences between Data Input and Groups

Results of the two-way ANOVA showed a statistically significant main effect of data input (F(1,78) =1854.44, *p*<0.001, partial η^2^ = 0.960) and group (F(1,78) =22.050, *p*<0.001, partial η^2^ = 0.220). Independent of the group, results of the pairwise-comparisons showed that prediction of activity across all deep brain regions was less accurate when using simulated fNIRS data than when using cortical fMRI data (mean difference = -0.654(95% CI, -0.684 to -0.624), FDR-corrected *p*<0.001). Independent of the data input, results of the pairwise-comparisons showed that prediction of activity across all deep brain regions was less accurate in the PTSD group than in the healthy control group (mean difference = -0.133 (95% CI, -0.190 to -0.077), FDR-corrected *p*<0.001). Follow-up paired sample t-tests showed higher prediction performance for cortical fMRI input compared to simulated fNIRS input for healthy controls (t(39) =27.995, FDR-corrected *p*<0.001, mean difference = 0.646 (95% CI, 0.599 to 0.692)) and individuals with PTSD (t(39) =33.511, FDR-corrected *p*<0.001, mean difference = 0.662 (95% CI, 0.622 to 0.702)). Further, we observed higher prediction performance in the healthy control group compared to individuals with PTSD when using cortical fMRI data (t(39) =8.117, FDR-corrected *p*<0.001, mean difference = 0.125 (95% CI, 0.094 to 0.156)) and when using simulated fNIRS data as input (t(39) =12.495, FDR-corrected *p*<0.001, mean difference = 0.141 (95% CI, 0.119 to 0.165)).

### 3.3 Descriptive Information from the Prediction Weight Maps (Figure 2 & 3)

The strongest predictors of amygdala activity based on cortical fMRI data (see **Figure 2**) were regions implicated in regulation of limbic activity, including parahippocampal gyrus, insular cortex, and frontal orbital cortex; as well as regions implicated in face processing and vision, including the fusiform gyrus and lingual gyrus (normalized prediction weights >0.200). These regions were the top predictors of amygdala activity in both groups, and were all positive in valence, suggesting that activation within these regions was positively associated with amygdala activation. These same regions were also among the strongest positive predictors of hippocampus activity, as was the posterior cingulate gyrus (normalized prediction weights >0.230). In contrast, weight maps for amygdala and hippocampus activity inference based on simulated fNIRS data (see **Figure 3**) showed that the middle temporal gyrus (MTG), the temporoparietal junction (TPJ), and inferior visual association cortex had the strongest positive prediction weights (normalized prediction weights >0.205).

## 4 Discussion

In this study, we demonstrated the feasibility of inferring activity in subcortical regions that are central to the pathophysiology of PTSD (i.e., amygdala and hippocampus) using cortical fMRI and simulated fNIRS activity in a group of adolescents with PTSD (*N*=20) and an age-matched healthy control group (*N*=20). We tested different prediction models, including linear regression, a MLP Neutral Network, and a KKN model. Results of the cortical fMRI data analyses showed high prediction performances for the bilateral amygdala (*r>*0.91) and bilateral hippocampus (*r>*0.95) in both groups. Using fNIRS simulated data, relatively high prediction performance across all four deep brain regions was maintained in healthy controls (*r>*0.79) and youths with PTSD (*r>*0.75). Linear regression consistently outperformed the other prediction models for the cortical fMRI prediction (both groups) and simulated fNIRS prediction in healthy controls. In contrast, for simulated fNIRS predictions in youths with PTSD, the MLP neural network demonstrated the highest performance.

Numerous prior studies have mapped the anatomical and functional connectivity between subcortical regions (e.g., amygdala and hippocampus) and surface regions of the cortex (Bullmore and Sporns 2009; Power et al. 2011). To create a “gold-standard” benchmark, we first validated the feasibility of inferring fMRI deep brain activity of the bilateral amygdala and bilateral hippocampus based on fMRI activity from 96 cortical regions (as defined by the Harvard Oxford Cortical Atlas [Frazier et al. 2005; Desikan et al. 2006; Makris et al. 2006; Goldstein et al. 2007]). For healthy controls, results showed overall high prediction performance across all four deep brain regions (average of *r*=0.94 across all deep brain regions). These values exceed the results from the previous deep brain inference study in healthy adults (average *r*=0.76 across all deep brain regions, [Liu et al. 2015]). One potential explanation for the increase in prediction performance in the current analysis compared to the previous study is that we used a different method to define cortical fMRI regions and we trained the models using more cortical fMRI data. The high prediction performances observed in this study appear to be robust to potential “noise” possibly introduced by the considerable age range in both groups and related variability in functional connectivity related to brain development (Blakemore et al. 2010; Goddings et al. 2014; Vijayakumar et al. 2018). In addition, we train/test-splitted the data subject-wise (70%/30%) to ensure generalizability across different subjects within the same group. This prediction approach generally performs more poorly than within-individual train/test-splitting. The high prediction performances observed in this study are therefore promising results. Compared to healthy controls, we observed a minimal reduction in prediction performance in the PTSD group for cortical fMRI data input (average reduction: *r*=0.012). Nevertheless, the prediction performance was still very high in the PTSD group (bilateral amygdala r*>*0.91; bilateral hippocampus r*>*0.95). With respect to our hypothesis, we can confirm the feasibility of the deep brain inference method in both youths with PTSD and healthy controls on the basis of cortical fMRI input data.

The weight maps in **Figure 2**, showed that fMRI data in cortical regions that are involved in emotion processing and regulation (e.g., insular cortex, parahippocampal gyrus and inferior frontal gyrus; [Ochsner et al. 2012; Frank et al. 2014]) and in face processing (e.g., fusiform gyrus; [93, 94]) were heavily involved in predicting deep brain activity of the bilateral amygdala. For activity prediction of the bilateral hippocampus, the fMRI data weight maps show the strongest associations with the posterior cingulate cortex – a hub of the default mode network [95, 96] and a region associated with processing emotion in the context of autobiographical memory [106]. These findings are in line with recent evidence showing connectivity between these regions both at rest and during affective states [97, 98]. Differences in prediction performances between the groups is likely attributed to aberrant activity and connectivity patterns in PTSD [104,105]. For example, **Figure 3** showed that the PTSD group had consistently weaker prediction weights in the left dorsolateral prefrontal cortex (dlPFC) compared to healthy controls (across all four subcortical target regions). One interpretation of this finding is that PTSD is characterized by relatively weaker functional connectivity between the dlPFC and subcortical regions. Interestingly, previous studies have identified dysregulated brain functioning during negative emotion processing and regulation in the left dlPFC [22, 26, 29, 50]. The dlPFC is thought to play a key role in emotion through regulation of activity in the limbic system [80, 81]. The negative weights within the left dlPFC highlight the reciprocal relation between this region and activity in the limbic system (i.e., amygdala and hippocampus). In this context, it is worth noting that there is debate about whether aberrant functional connectivity in PTSD reflects alterations in top-down regulation of the limbic system (e.g., amygdala) via the prefrontal cortex [34-39] or altered reciprocal regulation between these regions [108,109]. It is unclear whether the temporal resolution of fMRI (e.g., 1-2 seconds) is suitable for studying these dynamic and directional connections. However, inferring deep brain activity with prefrontal fNIRS activity, which provides much higher temporal resolution than fMRI (e.g., *>*10Hz), could advance our understanding of fronto-limbic circuit dynamics. We believe that this research would improve our understanding of the brain mechanisms underlying PTSD, as well as other disorders that involve fronto-limbic alterations.

Compared to cortical fMRI prediction in healthy controls, results of simulated fNIRS data showed overall lower prediction performances for healthy controls (average reduction: *r*=0.120). This decline in performance is perhaps unsurprising given that simulated fNIRS data provide less extensive coverage of the cortex than the cortical fMRI data input. Specifically, the number of voxels in simulated fNIRS data was only 19.85% of number of voxels of cortical fMRI data. We therefore believe that the relatively high prediction performances (r*>*0.79) found in this study are promising. Specifically, these prediction performance values exceed the results from the previous deep brain inference study in healthy adults (i.e., the top 15% of predictions using fNIRS signals achieved an accuracy of 0.70, [40]). One potential explanation for our increased prediction performance is that we utilized simulated fNIRS data across the entire “fNIRS-accessible” cortex, whereas the prior study utilized fNIRS data from prefrontal regions only [40]. These results suggest that future research on deep brain inference methods should consider assessment across the entire cortex rather than focusing on distinct regions of interest only.

In line with the cortical fMRI findings, we also observed slight reductions in prediction performance for the PTSD cohort (average reduction: *r*=0.047), which may be due to aberrant cortical activity and connectivity patterns in PTSD. More specifically, we consistently observed increased weights in the bilateral middle temporal gyrus (MTG) and left temporoparietal junction (TPJ) for the PTSD group across all four deep brain regions. Prior study findings suggest that the MTG and TPJ are involved in the general perceptual recognition of facial expressions [82, 83]. Increased activation of the MTG and TPJ have also been linked to dissociation and depersonalization [50, 84, 85], both key symptoms in PTSD [86–89]. The higher weights in the MTG and TPJ suggests that youths with PTSD recruited more neural resources to process emotional faces (e.g., evaluate, regulate affective response, dissociate or distance from). Interestingly, in contrast to the cortical fMRI findings, we did not observe high negative weights in the left dlPFC. These findings could suggest that activity in deeper portions of the left dlPFC are involved in fronto-limbic circuits. In this context, it is important to highlight that we chose a conservative approach to simulate the penetration depth for the fNIRS sensitivity maps. Actual fNIRS data from youths with PTSD demonstrate that fNIRS can detect left dlPFC activity during the facial expression task [50]. Future research is necessary to extend this proof-of-concept to concurrent fNIRS-fMRI assessment to demonstrate the feasibility of the deep brain inference method with actual fNIRS data. With respect to our hypothesis, we can confirm the feasibility of the deep brain inference method in both youths with PTSD and healthy controls on the basis of simulated fNIRS input data.

Linear regression (without rolling window) provided the best prediction performance in both groups when using fMRI cortical data, and in healthy controls when using simulated fNIRS cortical data. In contrast, the MLP neural network was the best performing model in the PTSD group when using simulated fNIRS data. *With respect to level of linearity*, these findings suggest that for those cases that achieved highest prediction performance with linear regression (without rolling window), the underlying problem setting was sufficiently linear. In contrast, MLP Neural Networks are capable of expressing non-linear relationships in the features. Their better performance for the simulated fNIRS input data in youths with PTSD compared to healthy controls suggests that the change in input data made predictive relationships more non-linear. *With respect to measurement noise*, the considerably smaller number of voxels per cortical region for the simulated fNIRS data increased the variance of the time-averaged signal per cortical region. Findings indicate that the use of a dropout layer for the MLP neural network helped to prevent overfitting to noise. The underperformance of the KNN Model compared to linear regression (without rolling window) and MLP neural network for all four groups is an additional indicator of the noisiness of the input data. In KNN models, noise is treated as part of the underlying dynamical model and increased noise can therefore cause overfitting (i.e., reduction in prediction performance). As we tested multiple values for *K*, we achieved the best performance for *K* = 23 for the control group and *K* = 15 for the PTSD group, both with cortical fMRI data as input. For the experiments, with simulated fNIRS data as input, *K* = 24 for the controls group and *K* = 10 for the PTSD group resulted in the best correlation values. These high values indicate that the training data is noisy which leads to oversmoothing for the predictions losing important information given the limited size of the dataset. *With respect to time-dependency*, we decided to include linear regression with a rolling window of size 3 (i.e., time step *t, t* − 1, and *t* − 2) to gain additional insights on the time scale of the underlying cortical-deep brain connectivity pattern. Results demonstrated the linear regression with rolling window of size 3 performed worst across all different tests. These results indicate that the integration of additional features (i.e., data from previous timesteps) effectively decreased the prediction accuracy leading to the conclusion that values from previous timesteps contain limited valuable information for the predictions. In summary, these findings suggest that future studies can benefit from utilizing both linear regression and MLP neural networks.

We note two limitations. First, our sample size is limited (i.e., *N*=20 in each group). A larger sample size could further increase the performances of the prediction models. Nevertheless, it is important to reiterate that we achieved high prediction performance, despite the relatively small sample size. A second limitation of this study is that we utilized simulated fNIRS data. Given the high cost of MRI scanning, we decided to conduct this proof-of-concept study before advancing to concurrent fNIRS-fMRI data collection. Our findings suggest that future fNIRS studies utilizing deep brain inference methods should attempt to increase their coverage of the cortex, ideally with high density coverage. A reduction in optodes (e.g., 10-20 positions) could potentially lead to reduced prediction performances. Additional processing steps will be required to temporally coordinate data signals from fNIRS (*>*10Hz) and fMRI (*<*2Hz). To temporally coordinate fNIRS and fMRI signals, researchers have established procedures to process latencies based on disparate sampling frequencies [40, 41, 90, 91].

In conclusion, this study provides further support about the potential of inferring fMRI deep brain activity with fNIRS. We demonstrated the feasibility of inferring fMRI deep brain activity (i.e., amygdala and hippocampus) based on cortical fMRI and simulated fNIRS activity in youths with PTSD and healthy controls. The overall high performances of these prediction models is particularly promising given some of our methodological constraints: we utilized a relatively small dataset, with youths undergoing brain development, and train/test-splitted between individuals (70/30). Further development of the deep brain inference method could lead to important advances in PTSD research and clinical applications. Specifically, fNIRS-derived deep brain biomarkers could inform clinical diagnostic and optimal treatment planning, potentially at a more scalable-level than what is feasible with MRI.

## Data Availability

Data will be available upon peer-reviewed acceptance of the manuscript.

## Acknowledgement

This work was supported by a K01 award from the National Institute of Mental Health to Dr. Garrett (NIMH K01 MH09776901A1).

## Author Statement

Stephanie Balters (Conceptualization, Formal analysis, Methodology, Writing – original draft, review and editing), Marc Schlichting (Formal analysis, Methodology, Writing – original draft, review and editing), Lara Foland-Ross (Data curation), Sabrina Brigadoi (Formal analysis), Jonas G. Miller (Writing – review and editing draft), Amy S. Garrett (Data curation, Funding), Mykel J. Kochenderfer (Methodology, Supervision, Writing - review editing), Allan L. Reiss (Conceptualization, Methodology, Supervision, Writing - review editing)

## Footnotes

1 ^1^ For an introduction to fNIRS, we refer the interested reader to [43, 44]

## References

[1] William E Copeland, Gordon Keeler, Adrian Angold, and E Jane Costello. Traumatic events and posttraumatic stress in childhood. Archives of general psychiatry, 64(5):577–584, 2007.

[2] Rose M Giaconia, Helen Z Reinherz, Amy B Silverman, Bilge Pakiz, Abbie K Frost, and Elaine Cohen. Traumas and posttraumatic stress disorder in a community population of older adolescents. Journal of the American Academy of Child & Adolescent Psychiatry, 34(10):1369–1380, 1995.

[3] Eva Alisic, Alyson K Zalta, Floryt Van Wesel, Sadie E Larsen, Gertrud S Hafstad, Katayun Hassanpour, and Geert E Smid. Rates of post-traumatic stress disorder in trauma-exposed children and adolescents: metaanalysis. The British Journal of Psychiatry, 204(5):335–340, 2014.

[4] Alexandra Macdonald, Carla Kmett Danielson, Heidi S Resnick, Benjamin E Saunders, and Dean G Kilpatrick. Ptsd and comorbid disorders in a representative sample of adolescents: The risk associated with multiple exposures to potentially traumatic events. Child abuse & neglect, 2010.

[5] Carl F Weems, Bethany H McCurdy, and Mikaela D Scozzafava. Toward a developmental model of continuity and change in ptsd symptoms following exposure to traumatic and adverse experiences. Journal of Child & Adolescent Trauma, pages 1–12, 2021.

[6] Carl F Weems, Justin D Russell, Ryan J Herringa, and Victor G Carrion. Translating the neuroscience of adverse childhood experiences to inform policy and foster population-level resilience. American Psychologist, 76(2):188, 2021.

[7] M Aebi, Meichun Mohler-Kuo, S Barra, U Schnyder, T Maier, and MA Landolt. Posttraumatic stress and youth violence perpetration: A population-based cross-sectional study. European psychiatry, 40:88–95, 2017.

[8] Susan Yoon, Stacey Steigerwald, Megan R Holmes, and Adam T Perzynski. Children’s exposure to violence: The underlying effect of posttraumatic stress symptoms on behavior problems. Journal of traumatic Stress, 29(1):72–79, 2016.

[9] Stephanie Malarbi, Hisham Motkal Abu-Rayya, F Muscara, and R Stargatt. Neuropsychological functioning of childhood trauma and post-traumatic stress disorder: A meta-analysis. Neuroscience & Biobehavioral Reviews, 72:68–86, 2017.

[10] Victor G Carrión and Carl Weems. Neuroscience of pediatric PTSD. Oxford University Press, 2017.

[11] Cohen, J. A., Deblinger, E., & Mannarino, A. P. (2018). Trauma-focused cognitive behavioral therapy for children and families. Psychotherapy Research, 28(1), 47–57.

[12] Christine Anne Rabinak, Mike Angstadt, Robert C Welsh, Amy Kennedy, Mark Lyubkin, Brian Martis, and K Luan Phan. Altered amygdala resting-state functional connectivity in post-traumatic stress disorder. Frontiers in psychiatry, 2:62, 2011.

[13] Feng Chen, Jun Ke, Rongfeng Qi, Qiang Xu, Yuan Zhong, Tao Liu, Jianjun Li, Li Zhang, and Guangming Lu. Increased inhibition of the amygdala by the mpfc may reflect a resilience factor in post-traumatic stress disorder: a resting-state fmri granger causality analysis. Frontiers in psychiatry, 9:516, 2018.

[14] Qiyong Gong, Lingjiang Li, Mingying Du, William Pettersson-Yeo, Nicolas Crossley, Xun Yang, Jing Li, Xiaoqi Huang, and Andrea Mechelli. Quantitative prediction of individual psychopathology in trauma survivors using resting-state fmri. Neuropsychopharmacology, 39(3):681–687, 2014.

[15] Jennifer S Stevens, Ye Ji Kim, Isaac R Galatzer-Levy, Renuka Reddy, Timothy D Ely, Charles B Nemeroff, Lauren A Hudak, Tanja Jovanovic, Barbara O Rothbaum, and Kerry J Ressler. Amygdala reactivity and anterior cingulate habituation predict posttraumatic stress disorder symptom maintenance after acute civilian trauma. Biological psychiatry, 81(12):1023–1029, 2017.

[16] Ye Ji Kim, Sanne JH van Rooij, Timothy D Ely, Negar Fani, Kerry J Ressler, Tanja Jovanovic, and Jennifer S Stevens. Association between posttraumatic stress disorder severity and amygdala habituation to fearful stimuli. Depression and anxiety, 36(7):647–658, 2019.

[17] Masaya Misaki, Raquel Phillips, Vadim Zotev, Chung-Ki Wong, Brent E Wurfel, Frank Krueger, Matthew Feldner, and Jerzy Bodurka. Connectome-wide investigation of altered resting-state functional connectivity in war veterans with and without posttraumatic stress disorder. NeuroImage: Clinical, 17:285–296, 2018.

[18] Xin Wang, Hong Xie, Andrew S Cotton, Elizabeth R Duval, Marijo B Tamburrino, Kristopher R Brickman, Jon D Elhai, S Shaun Ho, Samuel A McLean, Eric J Ferguson, et al. Preliminary study of acute changes in emotion processing in trauma survivors with ptsd symptoms. PLoS One, 11(7):e0159065, 2016.

[19] Jacklynn M Fitzgerald, Emily L Belleau, Tara A Miskovich, Walker S Pedersen, and Christine L Larson. Multivoxel pattern analysis of amygdala functional connectivity at rest predicts variability in posttraumatic stress severity. Brain and Behavior, 10(8):e01707, 2020.

[20] Richard A Bryant, Thomas Williamson, May Erlinger, Kim L Felmingham, Gin Malhi, Mark Hinton, Leanne Williams, and Mayuresh S Korgaonkar. Neural activity during response inhibition associated with improvement of dysphoric symptoms of ptsd after trauma-focused psychotherapy—an eeg-fmri study. Translational psychiatry, 11(1):1–10, 2021.

[21] Reinoud Kaldewaij, Saskia BJ Koch, Mahur M Hashemi, Wei Zhang, Floris Klumpers, and Karin Roelofs. Anterior prefrontal brain activity during emotion control predicts resilience to post-traumatic stress symptoms. Nature human behaviour, 5(8):1055–1064, 2021.

[22] Gregory A Fonzo, Madeleine S Goodkind, Desmond J Oathes, Yevgeniya V Zaiko, Meredith Harvey, Kathy K Peng, M Elizabeth Weiss, Allison L Thompson, Sanno E Zack, Steven E Lindley, et al. Ptsd psychotherapy outcome predicted by brain activation during emotional reactivity and regulation. American Journal of Psychiatry, 174(12):1163–1174, 2017.

[23] Sonalee A Joshi, Elizabeth R Duval, Jony Sheynin, Anthony P King, K Luan Phan, Brian Martis, Katherine E Porter, Israel Liberzon, and Sheila AM Rauch. Neural correlates of emotional reactivity and regulation associated with treatment response in a randomized clinical trial for posttraumatic stress disorder. Psychiatry Research: Neuroimaging, 299:111062, 2020.

[24] Shelly Sheynin, Lior Wolf, Ziv Ben-Zion, Jony Sheynin, Shira Reznik, Jackob Nimrod Keynan, Roee Admon, Arieh Shalev, Talma Hendler, and Israel Liberzon. Deep learning model of fmri connectivity predicts ptsd symptom trajectories in recent trauma survivors. Neuroimage, 238:118242, 2021.

[25] Emily L Belleau, Lauren E Ehret, Jessica L Hanson, Karen J Brasel, Christine L Larson, and Terri A deRoon Cassini. Amygdala functional connectivity in the acute aftermath of trauma prospectively predicts severity of posttraumatic stress symptoms. Neurobiology of Stress, 12:100217, 2020.

[26] Amy S Garrett, Victor Carrion, Hilit Kletter, Asya Karchemskiy, Carl F Weems, and Allan Reiss. Brain activation to facial expressions in youth with ptsd symptoms. Depression and anxiety, 29(5):449–459, 2012.

[27] Taylor J Keding and Ryan J Herringa. Paradoxical prefrontal–amygdala recruitment to angry and happy expressions in pediatric posttraumatic stress disorder. Neuropsychopharmacology, 41(12):2903–2912, 2016.

[28] Victor G Carrion, Brian W Haas, Amy Garrett, Suzan Song, and Allan L Reiss. Reduced hippocampal activity in youth with posttraumatic stress symptoms: an fmri study. Journal of pediatric psychology, 35(5):559–569, 2010.

[29] Amy Garrett, Judith A Cohen, Sanno Zack, Victor Carrion, Booil Jo, Joseph Blader, Alexis Rodriguez, Thomas J Vanasse, Allan L Reiss, and W Stewart Agras. Longitudinal changes in brain function associated with symptom improvement in youth with ptsd. Journal of psychiatric research, 114:161–169, 2019.

[30] JM Cisler, BA Sigel, JS Steele, S Smitherman, K Vanderzee, J Pemberton, TL Kramer, and CD Kilts. Changes in functional connectivity of the amygdala during cognitive reappraisal predict symptom reduction during traumafocused cognitive–behavioral therapy among adolescent girls with post-traumatic stress disorder. Psychological medicine, 46(14):3013–3023, 2016.

[31] Alexander J Shackman, Tim V Salomons, Heleen A Slagter, Andrew S Fox, Jameel J Winter, and Richard J Davidson. The integration of negative affect, pain and cognitive control in the cingulate cortex. Nature Reviews Neuroscience, 12(3):154–167, 2011.

[32] Francis L Stevens, Robin A Hurley, and Katherine H Taber. Anterior cingulate cortex: unique role in cognition and emotion. The Journal of neuropsychiatry and clinical neurosciences, 23(2):121–125, 2011.

[33] Sarah J Banks, Kamryn T Eddy, Mike Angstadt, Pradeep J Nathan, and K Luan Phan. Amygdala–frontal connectivity during emotion regulation. Social cognitive and affective neuroscience, 2(4):303–312, 2007.

[34] M Alexandra Kredlow, Robert J Fenster, Emma S Laurent, Kerry J Ressler, and Elizabeth A Phelps. Prefrontal cortex, amygdala, and threat processing: implications for ptsd. Neuropsychopharmacology, 47(1):247–259, 2022.

[35] Wen-Hua Zhang, Jun-Yu Zhang, Andrew Holmes, and Bing-Xing Pan. Amygdala circuit substrates for stress adaptation and adversity. Biological Psychiatry, 89(9):847–856, 2021.

[36] Saskia BJ Koch, Mirjam van Zuiden, Laura Nawijn, Jessie L Frijling, Dick J Veltman, and Miranda Olff. Aberrant resting-state brain activity in posttraumatic stress disorder: A meta-analysis and systematic review. Depression and anxiety, 33(7):592–605, 2016.

[37] Rebecca K Sripada, Anthony P King, Sarah N Garfinkel, Xin Wang, Chandra S Sripada, Robert C Welsh, and Israel Liberzon. Altered resting-state amygdala functional connectivity in men with posttraumatic stress disorder. Journal of Psychiatry and Neuroscience, 37(4):241–249, 2012.

[38] Nathaniel G Harnett, Adam M Goodman, and David C Knight. Ptsd-related neuroimaging abnormalities in brain function, structure, and biochemistry. Experimental neurology, 330:113331, 2020.

[39] Ryan J Herringa. Trauma, ptsd, and the developing brain. Current psychiatry reports, 19(10):1–9, 2017.

[40] Ning Liu, Xu Cui, Daniel M Bryant, Gary H Glover, and Allan L Reiss. Inferring deep-brain activity from cortical activity using functional near-infrared spectroscopy. Biomedical optics express, 6(3):1074–1089, 2015.

[41] Xu Cui, Signe Bray, and Allan L Reiss. Functional near infrared spectroscopy (nirs) signal improvement based on negative correlation between oxygenated and deoxygenated hemoglobin dynamics. Neuroimage, 49(4):3039–3046, 2010.

[42] Felix Scholkmann, Stefan Kleiser, Andreas Jaakko Metz, Raphael Zimmermann, Juan Mata Pavia, Ursula Wolf, and Martin Wolf. A review on continuous wave functional near-infrared spectroscopy and imaging instrumentation and methodology. Neuroimage, 85:6–27, 2014.

[43] Xu Cui, Signe Bray, Daniel M Bryant, Gary H Glover, and Allan L Reiss. A quantitative comparison of nirs and fmri across multiple cognitive tasks. Neuroimage, 54(4):2808–2821, 2011.

[44] Gary Strangman, Joseph P Culver, John H Thompson, and David A Boas. A quantitative comparison of simultaneous bold fmri and nirs recordings during functional brain activation. Neuroimage, 17(2):719–731, 2002.

[45] Jonathan D Power, Alexander L Cohen, Steven M Nelson, Gagan S Wig, Kelly Anne Barnes, Jessica A Church, Alecia C Vogel, Timothy O Laumann, Fran M Miezin, Bradley L Schlaggar, et al. Functional network organization of the human brain. Neuron, 72(4):665–678, 2011.

[46] Ed Bullmore and Olaf Sporns. Complex brain networks: graph theoretical analysis of structural and functional systems. Nature reviews neuroscience, 10(3):186–198, 2009.

[47] Sarah-Jayne Blakemore, Stephanie Burnett, and Ronald E Dahl. The role of puberty in the developing adolescent brain. Human brain mapping, 31(6):926–933, 2010.

[48] Anne-Lise Goddings, Kathryn L Mills, Liv S Clasen, Jay N Giedd, Russell M Viner, and Sarah-Jayne Blakemore. The influence of puberty on subcortical brain development. Neuroimage, 88:242–251, 2014.

[49] Nandita Vijayakumar, Zdena Op de Macks, Elizabeth A Shirtcliff, and Jennifer H Pfeifer. Puberty and the human brain: Insights into adolescent development. Neuroscience & Biobehavioral Reviews, 92:417–436, 2018.

[50] Stephanie Balters, Rihui Li, Flint M Espil, Aaron Piccirilli, Ning Liu, Andrew Gundran, Victor G Carrion, Carl F Weems, Judith A Cohen, and Allan L Reiss. Functional near-infrared spectroscopy brain imaging predicts symptom severity in youth exposed to traumatic stress. Journal of psychiatric research, 144:494–502, 2021.

[51] Nikos Makris, Jill M Goldstein, David Kennedy, Steven M Hodge, Verne S Caviness, Stephen V Faraone, Ming T Tsuang, and Larry J Seidman. Decreased volume of left and total anterior insular lobule in schizophrenia. Schizophrenia research, 83(2-3):155–171, 2006.

[52] Jean A Frazier, Sufen Chiu, Janis L Breeze, Nikos Makris, Nicholas Lange, David N Kennedy, Martha R Herbert, Eileen K Bent, Vamsi K Koneru, Megan E Dieterich, et al. Structural brain magnetic resonance imaging of limbic and thalamic volumes in pediatric bipolar disorder. American Journal of Psychiatry, 162(7):1256–1265, 2005.

[53] Rahul S Desikan, Florent Ségonne, Bruce Fischl, Brian T Quinn, Bradford C Dickerson, Deborah Blacker, Randy L Buckner, Anders M Dale, R Paul Maguire, Bradley T Hyman, et al. An automated labeling system for subdividing the human cerebral cortex on mri scans into gyral based regions of interest. Neuroimage, 31(3):968–980, 2006.

[54] Jill M Goldstein, Larry J Seidman, Nikos Makris, Todd Ahern, Liam M O’Brien, Verne S Caviness Jr, David N Kennedy, Stephen V Faraone, and Ming T Tsuang. Hypothalamic abnormalities in schizophrenia: sex effects and genetic vulnerability. Biological psychiatry, 61(8):935–945, 2007.

[55] Sabrina Brigadoi and Robert J Cooper. How short is short? optimum source–detector distance for shortseparation channels in functional near-infrared spectroscopy. Neurophotonics, 2(2):025005, 2015.

[56] Joan Kaufman, Boris Birmaher, David Brent, UMA Rao, Cynthia Flynn, Paula Moreci, Douglas Williamson, and Neal Ryan. Schedule for affective disorders and schizophrenia for school-age children-present and lifetime version (k-sads-pl): initial reliability and validity data. Journal of the American Academy of Child & Adolescent Psychiatry, 36(7):980–988, 1997.

[57] Alan M Steinberg, Melissa J Brymer, Soeun Kim, Ernestine C Briggs, Chandra Ghosh Ippen, Sarah A Ostrowski, Kevin J Gully, and Robert S Pynoos. Psychometric properties of the ucla ptsd reaction index: Part i. Journal of traumatic stress, 26(1):1–9, 2013.

[58] Dean Sabatinelli, Erica E Fortune, Qingyang Li, Aisha Siddiqui, Cynthia Krafft, William T Oliver, Stefanie Beck, and Joshua Jeffries. Emotional perception: meta-analyses of face and natural scene processing. Neuroimage, 54(3):2524–2533, 2011.

[59] Gary H Glover and Christine S Law. Spiral-in/out bold fmri for increased snr and reduced susceptibility artifacts. Magnetic Resonance in Medicine: An Official Journal of the International Society for Magnetic Resonance in Medicine, 46(3):515–522, 2001.

[60] Song Lai and Gary H Glover. Three-dimensional spiral fmri technique: a comparison with 2d spiral acquisition. Magnetic resonance in medicine, 39(1):68–78, 1998.

[61] Jongho Lee, Michael Lustig, Dong-hyun Kim, and John M Pauly. Improved shim method based on the minimization of the maximum off-resonance frequency for balanced steady-state free precession (bssfp). Magnetic Resonance in Medicine: An Official Journal of the International Society for Magnetic Resonance in Medicine, 61(6):1500–1506, 2009.

[62] Stephen M Smith. Fast robust automated brain extraction. Human brain mapping, 17(3):143–155, 2002.

[63] Mark Jenkinson, Peter Bannister, Michael Brady, and Stephen Smith. Improved optimization for the robust and accurate linear registration and motion correction of brain images. Neuroimage, 17(2):825–841, 2002.

[64] Mark Jenkinson and Stephen Smith. A global optimisation method for robust affine registration of brain images. Medical image analysis, 5(2):143–156, 2001.

[65] Vladimir Fonov, Alan C Evans, Kelly Botteron, C Robert Almli, Robert C McKinstry, D Louis Collins, Brain Development Cooperative Group, et al. Unbiased average age-appropriate atlases for pediatric studies. Neuroimage, 54(1):313–327, 2011.

[66] Katherine L Perdue and Solomon G Diamond. T1 magnetic resonance imaging head segmentation for diffuse optical tomography and electroencephalography. Journal of biomedical optics, 19(2):026011, 2014.

[67] Qianqian Fang and David A Boas. Tetrahedral mesh generation from volumetric binary and grayscale images. In 2009 IEEE international symposium on biomedical imaging: from nano to macro, pages 1142–1145. Ieee, 2009.

[68] Robert Oostenveld and Peter Praamstra. The five percent electrode system for high-resolution eeg and erp measurements. Clinical neurophysiology, 112(4):713–719, 2001.

[69] Sabrina Brigadoi, Domenico Salvagnin, Matteo Fischetti, and Robert J Cooper. Array designer: automated optimized array design for functional near-infrared spectroscopy. Neurophotonics, 5(3):035010, 2018.

[70] Martin Schweiger and Simon R Arridge. The toast++ software suite for forward and inverse modeling in optical tomography. Journal of biomedical optics, 19(4):040801, 2014.

[71] Frédéric Bevilacqua, Dominique Piguet, Pierre Marquet, Jeffrey D Gross, Bruce J Tromberg, and Christian Depeursinge. In vivo local determination of tissue optical properties: applications to human brain. Applied optics, 38(22):4939–4950, 1999.

[72] Anna Custo, William M Wells Iii, Alex H Barnett, Elizabeth MC Hillman, and David A Boas. Effective scattering coefficient of the cerebral spinal fluid in adult head models for diffuse optical imaging. Applied optics, 45(19):4747–4755, 2006.

[73] Gary Strangman, Maria Angela Franceschini, and David A Boas. Factors affecting the accuracy of nearinfrared spectroscopy concentration calculations for focal changes in oxygenation parameters. Neuroimage, 18(4):865–879, 2003.

[74] Diederik P Kingma and Jimmy Ba. Adam: A method for stochastic optimization. arXiv preprint 1412.6980, 2014.

[75] Nitish Srivastava, Geoffrey Hinton, Alex Krizhevsky, Ilya Sutskever, and Ruslan Salakhutdinov. Dropout: a simple way to prevent neural networks from overfitting. The journal of machine learning research, 15(1):1929–1958, 2014.

[76] Evelyn Fix. Discriminatory analysis: nonparametric discrimination, consistency properties, volume 1. USAF school of Aviation Medicine, 1985.

[77] Naomi S Altman. An introduction to kernel and nearest-neighbor nonparametric regression. The American Statistician, 46(3):175–185, 1992.

[78] Student. Probable error of a correlation coefficient. Biometrika, pages 302–310, 1908.

[79] Peter Henderson, Riashat Islam, Philip Bachman, Joelle Pineau, Doina Precup, and David Meger. Deep reinforcement learning that matters. In Proceedings of the AAAI conference on artificial intelligence, volume 32, 2018.

[80] Despina E Ganella, Marjolein EA Barendse, Jee H Kim, and Sarah Whittle. Prefrontal-amygdala connectivity and state anxiety during fear extinction recall in adolescents. Frontiers in human neuroscience, 11:587, 2017.

[81] Dylan G Gee. Sensitive periods of emotion regulation: influences of parental care on frontoamygdala circuitry and plasticity. 2016.

[82] S Petersen, J Baker, J Allman, R Andersen, G Essick, R Siegel, J Neal, D Winfield, and T Powell. Amblyopia: a multidisciplinary approach. proctor lecture. Invest Ophthalmol Vis Sci, 26(12):1704–1716, 1985.

[83] Ke Zhao, Jia Zhao, Ming Zhang, Qian Cui, and Xiaolan Fu. Neural responses to rapid facial expressions of fear and surprise. Frontiers in psychology, 8:761, 2017.

[84] Olaf Blanke, Christine Mohr, Christoph M Michel, Alvaro Pascual-Leone, Peter Brugger, Margitta Seeck, Theodor Landis, and Gregor Thut. Linking out-of-body experience and self processing to mental own-body imagery at the temporoparietal junction. Journal of Neuroscience, 25(3):550–557, 2005.

[85] Isadora Olive, Maria Densmore, Sherain Harricharan, Jean Théberge, Margaret C McKinnon, and Ruth Lanius. Superior colliculus resting state networks in post-traumatic stress disorder and its dissociative subtype. Human brain mapping, 39(1):563–574, 2018.

[86] Ruth A Lanius, Peter C Williamson, Kristine Boksman, Maria Densmore, Madhulika Gupta, Richard WJ Neufeld, Joseph S Gati, and Ravi S Menon. Brain activation during script-driven imagery induced dissociative responses in ptsd: a functional magnetic resonance imaging investigation. Biological psychiatry, 52(4):305–311, 2002.

[87] Ruth A Lanius, Peter C Williamson, Robyn L Bluhm, Maria Densmore, Kristine Boksman, Richard WJ Neufeld, Joseph S Gati, and Ravi S Menon. Functional connectivity of dissociative responses in posttraumatic stress disorder: a functional magnetic resonance imaging investigation. Biological psychiatry, 57(8):873–884, 2005.

[88] Kasia Kozlowska, Peter Walker, Loyola McLean, and Pascal Carrive. Fear and the defense cascade: clinical implications and management. Harvard review of psychiatry, 2015.

[89] Maggie Schauer and Thomas Elbert. Dissociation following traumatic stress. Zeitschrift für Psychologie/Journal of Psychology, 2015.

[90] Louis Gagnon, Meryem A Yücel, Mathieu Dehaes, Robert J Cooper, Katherine L Perdue, Juliette Selb, Theodore J Huppert, Richard D Hoge, and David A Boas. Quantification of the cortical contribution to the nirs signal over the motor cortex using concurrent nirs-fmri measurements. Neuroimage, 59(4):3933–3940, 2012.

[91] Ochsner, K. N., Silvers, J. A., & Buhle, J. T. (2012). Functional imaging studies of emotion regulation: a synthetic review and evolving model of the cognitive control of emotion. Annals of the new York Academy of Sciences, 1251(1), E1–E24

[92] Frank, D. W., Dewitt, M., Hudgens-Haney, M., Schaeffer, D. J., Ball, B. H., Schwarz, N. F., … & Sabatinelli, D. (2014). Emotion regulation: quantitative meta-analysis of functional activation and deactivation. Neuroscience & Biobehavioral Reviews, 45, 202–211.

[93] Rossion, B., Dricot, L., Devolder, A., Bodart, J. M., Crommelinck, M., De Gelder, B., & Zoontjes, R. (2000). Hemispheric asymmetries for whole-based and part-based face processing in the human fusiform gyrus. Journal of cognitive neuroscience, 12(5), 793–802.

[94] McCarthy, G., Puce, A., Gore, J. C., & Allison, T. (1997). Face-specific processing in the human fusiform gyrus. Journal of cognitive neuroscience, 9(5), 605–610.

[95] Chiara Bulgarelli, Anna Blasi, Simon Arridge, Samuel Powell, Carina CJM de Klerk, Victoria Southgate, Sabrina Brigadoi, William Penny, Sungho Tak, and Antonia Hamilton. Dynamic causal modelling on infant fnirs data: A validation study on a simultaneously recorded fnirs-fmri dataset. NeuroImage, 175:413–424, 2018.

[96] Raichle, M. E. (2015). The brain’s default mode network. Annual review of neuroscience, 38, 433–447.

[97] Alves, P. N., Foulon, C., Karolis, V., Bzdok, D., Margulies, D. S., Volle, E., & Thiebaut de Schotten, M. (2019). An improved neuroanatomical model of the default-mode network reconciles previous neuroimaging and neuropathological findings. Communications biology, 2(1), 1–14.

[98] Figueroa, C. A., Mocking, R. J., van Wingen, G., Martens, S., Ruhe, H. G., & Schene, A. H. (2017). Aberrant default-mode network-hippocampus connectivity after sad memory-recall in remitted-depression. Social cognitive and affective neuroscience, 12(11), 1803–1813.

[99] Morris, N. M. & Udry, J. R. (1980). Validation of a self-administered instrument to assess stage of adolescent development. Journal of youth and adolescence, 9(3), 271–280.

[100] Stuart Rusell and Peter Norvig (2020). Artificial Intelligence: A Modern Approach (4th ed.). Pearson.

[101] Hota, H. S., Handa, R., & Shrivas, A. K. (2017). Time series data prediction using sliding window based RBF neural network. International Journal of Computational Intelligence Research, 13(5), 1145–1156.

[102] Fix, E., & Hodges, J. L. (1989). Discriminatory analysis. Nonparametric discrimination: Consistency properties. International Statistical Review/Revue Internationale de Statistique, 57(3), 238–247.

[103] Altman, Naomi S. “An introduction to kernel and nearest-neighbor nonparametric regression.” The American Statistician46.3 (1992): 175–185.

[104] Bao, W., Gao, Y., Cao, L., Li, H., Liu, J., Liang, K., … & Huang, X. (2021). Alterations in large-scale functional networks in adult posttraumatic stress disorder: A systematic review and meta-analysis of resting-state functional connectivity studies. Neuroscience & Biobehavioral Reviews, 131, 1027–1036.

[105] Suarez-Jimenez, B., Albajes-Eizagirre, A., Lazarov, A., Zhu, X., Harrison, B. J., Radua, J., … & Fullana, M. A. (2020). Neural signatures of conditioning, extinction learning, and extinction recall in posttraumatic stress disorder: a meta-analysis of functional magnetic resonance imaging studies. Psychological medicine, 50(9), 1442–1451.

[106] Maddock, R. J., Garrett, A. S., & Buonocore, M. H. (2003). Posterior cingulate cortex activation by emotional words: fMRI evidence from a valence decision task. Human brain mapping, 18(1), 30–41.

[107] American Psychiatric Association. (2013). Diagnostic and statistical manual of mental disorders (5th ed.). https://doi.org/10.1176/appi.books.9780890425596

[108] Yizhar, O., & Klavir, O. (2018). Reciprocal amygdala–prefrontal interactions in learning. Current opinion in neurobiology, 52, 149–155.

[109] Chiba, T., Ide, K., Taylor, J. E., Boku, S., Toda, H., Kanazawa, T., … & Koizumi, A. (2021). A reciprocal inhibition model of alternations between under-/overemotional modulatory states in patients with PTSD. Molecular psychiatry, 26(9), 5023–5039.

